# Increased brain atrophy over 3 years in stroke survivors is linked to early post-stroke cognitive impairment

**DOI:** 10.1101/2021.08.03.21261461

**Authors:** Amy Brodtmann, Emilio Werden, Mohamed Salah Khlif, Laura J. Bird, Natalia Egorova, Michele Veldsman, Heath Pardoe, Graeme Jackson, Jennifer Bradshaw, David Darby, Toby Cumming, Leonid Churilov, Geoffrey Donnan

**Author notes:** **Corresponding author:** Amy Brodtmann, The Florey Institute of Neuroscience and Mental Health, Melbourne Brain Centre, 245 Burgundy Street, Heidelberg 3081, Australia.

## Abstract

**Background:** Stroke survivors are at high risk of dementia, associated with increasing age and vascular burden and with pre-existing cognitive impairment, older age. Brain atrophy patterns are recognised as signatures of neurodegenerative conditions, but the natural history of brain atrophy after stroke remains poorly described. We sought to determine whether stroke survivors who were cognitively normal at time of stroke had greater total brain (TBV) and hippocampal volume (HV) loss over 3 years than controls. We examined whether stroke survivors who were cognitively impaired (CI) at 3 months following their stroke had greater brain volume loss than cognitively normal (CN) stroke participants.

**Methods:** Cognition And Neocortical Volume After Stroke (CANVAS) study is a multi-centre cohort study of first-ever or recurrent adult ischaemic stroke participants compared to age- and sex-matched community controls. Participants were followed with MRI and cognitive assessments over 3 years and were free of a history of cognitive impairment or decline at inclusion. Our primary outcome measure was TBV change between 3 months and 3 years; secondary outcomes were TBV and HV change comparing CI and CN participants. We investigated associations between group status and brain volume change using a baseline-volume adjusted linear regression model with robust standard error.

**Results:** Ninety-three stroke (26 women, 66.7±12 years) and 39 control participants (15 women, 68.7±7 years) were available at 3 years. TBV loss in stroke patients was greater than controls: stroke mean (M)=20.3cm^3^±SD14.8cm^3^; controls M=14.2cm^3^±SD13.2cm^3^; (adjusted mean difference 7.88 95%CI [2.84,12.91] p-value=0.002). TBV decline was greater in those stroke participants who were cognitively impaired (M=30.7cm^3^; SD=14.2cm^3^) at 3 months (M=19.6cm^3^; SD=13.8cm^3^); (adjusted mean difference 10.42; 95%CI [3.04,17.80], p-value=0.006). No statistically significant differences in HV change were observed.

**Conclusions:** Ischaemic stroke survivors exhibit greater neurodegeneration compared to stroke-free controls. Brain atrophy is greater in stroke participants who were cognitively impaired people early after their stroke. Early cognitive impairment may predict greater subsequent atrophy, reflecting the combined impacts of stroke and vascular brain burden. Atrophy rates could serve as a useful biomarker for trials testing interventions to reduce post-stroke cognitive impairment.

## INTRODUCTION

Stroke care has been transformed in the last three decades by improved reperfusion treatments and rehabilitation therapies. Despite this, the global burden of stroke remains high, in part due to long-term cognitive impairments and increased risk of dementia^1^. Cumulative vascular risk factors increase dementia risk^2, 3^; conversely, good cardiovascular health reduces this risk^4^. Risk of stroke and of cognitive decline are also conflated: cognitive impairment can anticipate the development of incident stroke^5^ and be a manifestation of this risk^6^. One of the strongest risk factors for post-stroke cognitive impairment is a history of pre-stroke cognitive or functional decline^7^.

Brain atrophy precedes and predicts cognitive decline in many neurodegenerative syndromes, but the trajectories of brain volume loss and cognitive impairment in stroke survivors are poorly understood. These have been difficult to disentangle given relatively few post-stroke longitudinal cohort studies with high-quality imaging and cognitive data, often including people with pre-stroke cognitive impairment and dementia. Structural brain changes are already present at the time of stroke, with smaller hippocampal volumes and increased white matter hyperintensity (WMH) volumes^8^. Extensive white matter degeneration is evident at 3 months after stroke^9^, and hippocampal, thalamic and hemispheric atrophy continues over the first post-stroke year^10^. Brain atrophy rates are now being used as biomarkers for clinical trials in neurodegenerative diseases. They may serve as a useful tool to assess treatment response for interventions to reduce post-stroke cognitive impairment. An independent biomarker has several advantages over more traditionally used cognitive assessments, as atrophy rates are not dependent on language, educational attainment, culture, or socio-economic status: all factors known to determine performance on canonical cognitive tests.

We aimed to determine whether ischaemic stroke was associated with progressive neurodegeneration via a prospective, 3-year cohort study comparing participants with ischaemic stroke to their age- and sex-matched healthy controls. We included participants with no history of cognitive impairment or decline, cross-checked via interview with family members and treating health practitioners. In addition, we aimed to determine whether stroke survivors with cognitive impairment at 3 months exhibited greater brain atrophy over the subsequent 3 years compared to those who were cognitively normal. We hypothesized that stroke survivors would exhibit greater total brain volume loss than controls, and that those who were cognitively impaired at 3 months after stroke would exhibit greater brain volume loss than those who were cognitively normal.

## METHODS

We received ethics approval from Human Research Ethics Committees at each of the participating hospitals. All participants provided written consent in person in accordance with the Declaration of Helsinki.

The Cognition And Neocortical Volume Study (CANVAS) study is a multi-center cohort study including patients with ischaemic stroke and healthy controls and tested with serial MRI scanning and cognitive testing over 3 years^11^.

Patients were recruited between April 2012 and July 2015 from the Stroke Units at three university teaching hospitals in Melbourne, Australia: Austin Hospital, Box Hill Hospital, and Royal Melbourne Hospital. Control participants were recruited until September 2015 to optimize age- and sex-matching. All testing and brain imaging were done at the Melbourne Brain Centre, Austin Hospital campus.

### Participants

Stroke survivors with first-ever clinical or recurrent ischaemic stroke and no history of dementia were recruited within the first 3 months of their stroke. They were approached for recruitment as inpatients or via phone interview once discharged. They were included if they had an ischaemic stroke of any stroke subtype^7^, and excluded if there was pre-existing cognitive impairment (based on participant, primary care practitioner, and informant); could not have MRI (e.g., implanted metal, severe orthopnoea); had primary haemorrhagic stroke, TIA, or no clinically confirmed stroke; or were unlikely to survive 3 years due to severe medical illness.

Healthy age- and sex-matched control participants with no history of cognitive impairment were recruited from a pool of community participants who had previously volunteered in MRI studies and from local community groups. Spouses and age-appropriate family members of stroke participants were also approached to attempt socioeconomic matching. Inclusion and exclusion criteria were the same as for stroke patients, except for the stroke diagnosis.

### Measurements

#### Sociodemographic

We obtained information via interview: age, years of education, handedness, marital status, stroke and dementia family history, smoking pack years (number of cigarettes per day multiplied by years of smoking), alcohol consumption (low ≤14; high>standard drinks/week); history of depression, hypertension, type 2 diabetes mellitus (T2DM), hypercholesterolemia and atrial fibrillation (AF) given either via physician diagnosis or medication use. Body mass index (BMI; kg/m^2;^ low<25, high (≥25) was calculated using weight and height measurements.

#### Clinical

We obtained information of stroke side, use of thrombolysis, admission stroke severity (National Institute of Health Stroke Scale (NIHSS)) score, etiology^12^ and subtype (Oxfordshire criteria)^13^.We used the modified Rankin Scale^14^ (mRS) as an estimate of neurological disability and the Charlson Comorbidity Index (CCI)^15^, a validated co-morbidity score, as an estimate of general medical comorbidity, removing the stroke score for people with stroke.

Venous blood was drawn for *APOE* genotype determination on participants who consented to DNA analyses and storage. Individuals were categorised as *APOE* ε*4* carriers or noncarriers.

#### Imaging

All participants were scanned on the same MRI scanner which did not undergo significant hardware or software upgrades over the study period. Whole brain images were acquired on a single 3T Siemens Tim Trio Scanner with a 12-channel head coil (Siemens, Erlangen, Germany) (please see https://www.ahajournals.org/journal/str for details). All images were visually inspected for quality control before processing using automated pipelines and excluded if degraded by motion or other artefacts. Cortical reconstruction and volumetric segmentation on MPRAGE images were performed using the longitudinal stream (http://surfer.nmr.mgh.harvard.edu/fswiki/LongitudinalProcessing) in FreeSurfer V6.0^16^. Hippocampal results were based on averages of left and right hippocampal volumes.

Stroke lesions were traced by our imaging analyst (MSK) and cross-checked by a stroke neurologist (AB). Stroke lesion sites were also cross-checked with acute inpatient imaging, particularly their DWI lesion site on acute MRI where available. We did not adjust for stroke lesion volume as its association with post-stroke brain atrophy rates and cognition is not known^7^, and because it was fully correlated with stroke status (zero volume in controls); therefore, a feature not an artefact. We understand that there are vigorous arguments both for and against lesion volume adjustment. WMH probability maps were obtained from FLAIR images using the lesion prediction algorithm^17^ from the lesion segmentation toolbox included in SPM12.

#### Cognitive

We used the National Adult Reading Test (NART^18^) to estimate pre-morbid IQ and the Informant Questionnaire on Cognitive Decline in the Elderly (IQCODE-Short Form^19^) to estimate pre-morbid general cognitive functioning and to probe for the presence of preceding undiagnosed cognitive decline. Symptoms of anxiety and depression were examined at each session using the Generalized Anxiety Disorder-7 (GAD-7^20^) scale and Patient Health Questionnaire-9 (PHQ-9^21^) together with a clinical interview.

Neuropsychological testing was done in a single session allowing time for breaks. The cognitive testing protocol has been previously described^11^, and included: Hopkins Verbal Learning Test-Revised (HVLT-R^22^); Detection, Identification and One-Back computerised tests from the CogState Battery^23^; Rey-Osterrieth Complex Figure; Star Cancellation Task; Verbal Fluency Task (FAS and Animals); Trail-Making Test A and B; Digit Span and Digit-Symbol Tasks from the Weschler Adult Intelligence Scale-Third Edition (WAIS-III^24^); Token Test 16-item version; Boston Naming Test^25^; and Clock Drawing Test. Age-appropriate normative values (mean, standard deviation) were used where available to create z-scores for each cognitive task. A composite z-score was created for each cognitive domain by averaging z-scores across tasks: attention (focused attention, working memory, processing speed), executive function, memory, language, and visuospatial function.

A weighted Global Clinical Dementia Rating (CDR) Score^26^ was performed at the 3-year post-stroke assessment to examine for functional decline.

#### Cognitive Outcome Evaluation Committee

Participants were assigned their status of *normal cognition (CN), cognitively impaired (CI)*, or *dementia*, in these panel meetings. We also allowed the assignment of *unclassifiable* if the participant was unable to complete sufficient cognitive testing and/or no history was obtained from an informant.

A participant was assigned *cognitively normal* (CN) if z-scores in all cognitive domains were within accepted age- and years of education-adjusted norms and there was no evidence of functional decline due to cognitive impairment (i.e., activities of daily living (ADLs) were unaffected).

A participant was judged to be *CI* if (1) the z-score for at least one cognitive domain was lower than -1.5, and (2) there was no evidence of functional decline due to cognitive impairment (ADLs were unaffected).

A participant was judged to have *dementia* if (1) z-scores for two or more cognitive domains were lower than -1.5 and (2) there was evidence of functional decline (i.e., significant impact on activities of daily living between time-points due to cognitive impairment, as demonstrated by (a) discussions with next-of-kin and the Short-Form IQCODE, and/or (b) CDR score of 1 or greater.

### Outcome measures

#### Primary outcome

Our primary outcome measure was total brain volume (TBV) change between the 3-month and 3-year time-points compared between stroke patients and controls. TBV and CCI were chosen *a priori* as the adjustment covariates, as male sex is associated with larger heads^27^, and both medical co-morbidities^2^ and age^27^ are associated with brain atrophy.

#### Secondary outcomes

Secondary outcome 1 was TBV change between 3-months and 3-years comparing CN and CI stroke participants. We adjusted for TBV at 3-months, CCI scores, and years of education; the latter as it is correlated with cognitive performance and post-stroke dementia risk^7^.

Secondary outcome 2 was hippocampal volume (HV) change between 3-months and 3-years in stroke patients and controls with adjustments identical to primary outcome.

Secondary outcome 3 was the comparison of HV change between 3-months and 3-years comparing CN and CI stroke participants with adjustments identical to secondary outcome 1.

### Sample size calculation

We calculated our sample size to include sufficient participants for both the primary outcome and secondary outcome 1. We based our brain volume loss estimates on pilot^28^ and published data^27, 29^. We estimated 35 participants per group yielding power of 0.8 assuming two-sided alpha of 0.05. For the CI versus CN comparison, we estimated an expected prevalence of cognitive impairment of 30% at 3 months^7^. We used an ANCOVA method to estimate sample size for two samples with repeated measures (CN versus CI stroke in a ratio of 2:1), including a correlation score between baseline and follow-up. Using an alpha level of 0.05 (two-sided), power=0.8, and correlation=0.1, we estimated that 108 participants would be required (36 CI and 72 CN), with predicted 20% attrition due to death or non-participation, giving a total 135 stroke and 40 control participants.

### Statistical Analysis

Our statistical analysis plan was formulated prior to study database lock. All missing data were assumed to be missing-at-random following deliberations by the Cognitive Outcome Evaluation Committee prior to statistical analysis. After cognitive group allocation and analyses, we compared demographic and stroke characteristics in stroke patients who completed the 3-month and 3-year review sessions with those who withdrew from the study or were unable to attend study visits (Supplemental Tables II-III).

Patient characteristics were summarised as either means and SD, or medians and interquartile ranges (IQRs), for continuous variables; and counts (proportions) for categorical variables. They were compared between groups using t-test, Mann-Whitney test, or chi-square/Fisher Exact test depending on the distribution. We conducted our primary complete case analysis on the sub-sample of participants with data available at 3 months and 3 years.

We used a linear regression model with robust standard error estimation to investigate the primary outcome for the complete case analysis: TBV change between 3-months and 3-years as the dependent variable, group (stroke versus control) as independent variable, and TBV and CCI at 3 months as covariates. The same approach was used for Secondary Outcome 1 using CN versus CI group, and the inclusion of years of education as covariate.

We applied the same models for Secondary Outcomes 2 and 3 but used HV change as our dependent variable. We conducted further sensitivity analysis on the sub-sample of participants who had 3-month data irrespective of 3-year data availability under the assumption of missing-at-random. This sensitivity analysis was conducted using linear mixed-effects modelling with specific outcomes of interest as output variables; individual group, time-points, and multiplicative group-by-time interaction term as independent variables; adjustment covariates as specified above; and individual participants as random effects.

Analyses were done in Stata version 15.0 (StataCorp, College Station, TX, USA). Two-sided p-values less than 0.05 were regarded as indicative of statistical significance. No correction for multiplicity of comparisons to limit family-wise Type I error rate was undertaken.

## RESULTS

### Participants

Over the study period, 3037 stroke patients were admitted to the participating stroke units and screened for potential study recruitment; 2659 did not meet study criteria (i.e., haemorrhagic stroke, prior history of dementia, severe stroke making 3-year survival unlikely, etc.). Of the 378 potential participants, 203 declined study participation, 40 were not included for other reasons (e.g., rural or regional participant unable to travel to the scanner). We recruited 135 stroke participants (Supplemental Figure I).

Nine were not available at the 3-month study visit (two returned at 3 years). One-hundred and twenty-six stroke patients attended their testing session at 3 months: four had incomplete or non-evaluable MRI scans; one had incomplete cognitive testing. A total of 122 were available for primary outcome analyses and a total of 121 participants had complete data sets at 3 months (Table 1).

**Table 1.** Characteristics of stroke and control participants attending the 3-month assessment. Comparison of demographic, imaging and cognitive characteristics of stroke patients and healthy controls at 3 months. Significant p-values are in bold.

|  | Stroke |  | Control |  | *p* |
| --- | --- | --- | --- | --- | --- |
|  | N |  | N |  |  |
| Sociodemographic |  |  |  |  |  |
| Age, years, M±SD | 126 | 68.4(11.84) | 40 | 68.8(6.63) | 0.85 |
| Men, no.,(%) | 126 | 87(69.1%) | 40 | 25(62.5%) | 0.45 |
| Clinical |  |  |  |  |  |
| *APOE* ε4, no.,(%) | 112 | 20(17.9%) | 39 | 4(10.3%) | 0.32 |
| Depression, no. (%) | 126 | 12(9.5%) | 40 | 4(10%) | 1.0 |
| Charlson Comorbidity Scores<sup>15</sup>,<br>Md (Q1,Q3) | 126 | 3(2, 5) | 40 | 3(2, 3) | **0.03** |
| Hypertension, no.,(%) | 126 | 80(63.5%) | 40 | 17(42.5%) | **0.03** |
| Hypercholesterolaemia, no (%) | 126 | 57(45.2%) | 40 | 14(35%) | 0.28 |
| Type 2 diabetes mellitus,<br>No.,(%) | 126 | 32(25.4%) | 40 | 4(10%) | **0.047** |
| Atrial fibrillation, no.,(%) | 126 | 30(23.8%) | 40 | 1(2.5%) | **0.002** |
| Smoking, pack years,<br>Md (Q1,Q3) | 122 | 1(0, 17) | 40 | 0(0, 30) | **0.045** |
| Obesity, no.,(%) | 126 | 37(29.4%) | 40 | 7(17.5%) | 0.16 |
| Imaging |  |  |  |  |  |
| Days to scan (Md,Q1,Q3) | 122 | 92(85,112) | 40 | 94(87, 08) | 0.71 |
| Total brain volume (cm<sup>3</sup>,<br>M±SD) | 122 | 1079(123) | 40 | 1113(83) | 0.054 |
| Hippocampal volume (mm<sup>3</sup>,<br>M±SD) | 122 | 3862(490) | 40 | 4032(343) | **0.017** |
| Stroke lesion volume<br>(mm<sup>3</sup>,Md,Q1,Q3) | 122 | 1866(443,6869) | - | - | - |
| WMH Volume | 122 | 1866(443,6869) | 40 | 684(225,690) | **0.0001** |
| (mm<sup>3</sup>,Md,Q1,Q3) |  |  |  |  |  |
| **Cognitive** |  |  |  |  |  |
| Education, years,<br>Md (Q1,Q3) | 126 | 12(10,15) | 40 | 17(11,18) | **0.0005** |
| NART-FSIQ, Md (Q1,Q3) | 113 | 111<br>(111.5,124.9) | 40 | 120.9<br>(111.5,124.9) | **0.0002** |

At 3 years, 15 stroke participants had died, and 14 had moved interstate, were uncontactable, or withdrew for other reasons (n=29; Supplemental Tables I and III); 102 stroke participants returned for their study visit but nine had non-evaluable scans, meaning a total of 93 were included for the primary outcome analysis at 3 years; 92 were included for secondary outcomes 1 and 3 as one stroke participant had an incomplete cognitive assessment at 3 months (Table 2; Supplemental Tables II-III). Our attrition rate was 19% for stroke participants (102/126), in line with the 20% predicted for our analyses.

**Table 2.**
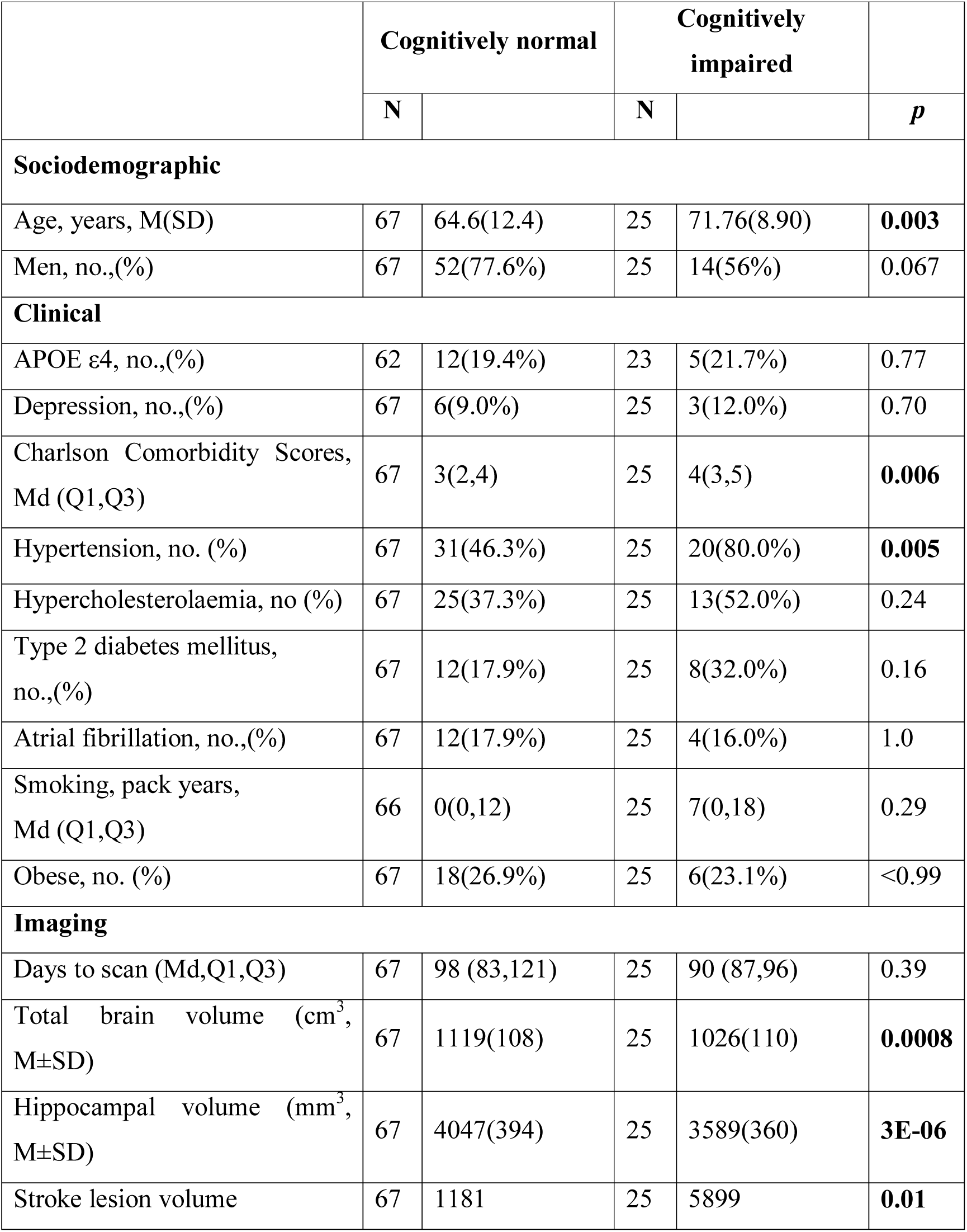
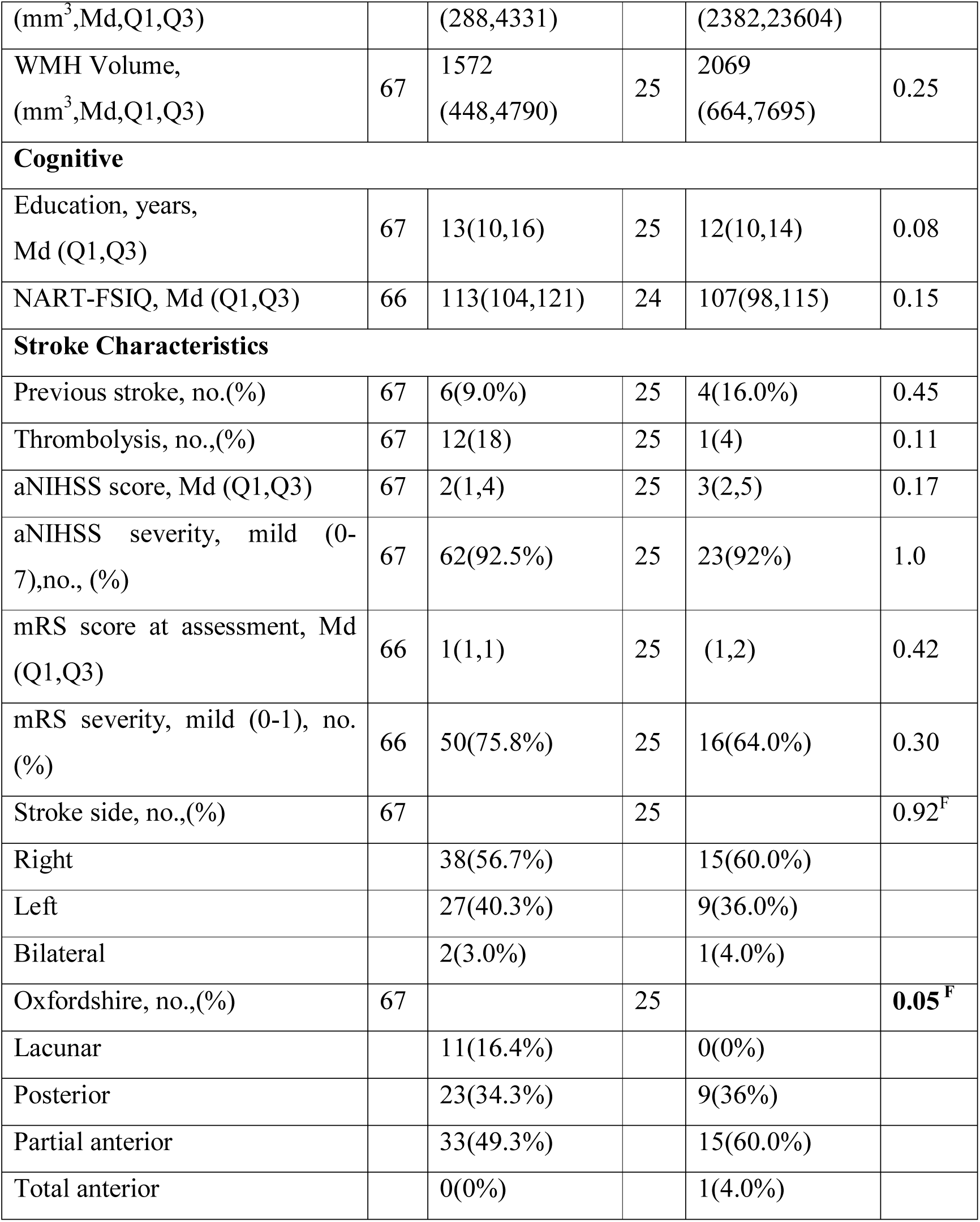
Characteristics of stroke participants who were cognitively normal versus cognitively impaired at 3-month visit. Stroke participant demographic, clinical, imaging, cognitive and stroke characteristics comparing CN versus CI at 3 months. ^F^=Fisher exact test value per category; aNIHSS=admission NIHSS. Significant p-values are in bold.

We examined for attendance and evaluable scan bias following database lock. Incomplete data sets were associated with obesity at both time-points, and with older age and history of hypertension at 3 years (Supplemental Tables II-III), but not with admission stroke severity. One-hundred and forty-six control participants were screened for study participation: 18 did not meet inclusion criteria, 22 declined participation and 66 did not respond to our invitation to participate. Forty control participants were included in the 3-month assessment and 39 were available for the 3-year assessment as one participant withdrew. Two participants had silent lesions noted on their research MRI scans, but no stroke diagnosis after clinical review, remaining in the study.

All control participants were cognitively normal at 3 months and 3 years. At 3 months, there were 80 CN and 41 CI stroke participants (Table 2). At 3 years, there were 73 CN, 16 CI and 4 dementia stroke participants, which included 67 CN and 25 CI assigned at the 3-month visit. Significantly more (p=0.016) 3-month CI participants died or did not attend their 3-year visit (34%) than CN (14%).

### Sociodemographic

No differences in age and sex were observed between groups at 3 months and 3 years.

### Clinical

Strokes were in all vascular territories (Supplemental Figure II). No differences in stroke characteristics were observed between stroke participants available at 3 months and 3 years (Supplemental Tables I-III).

At 3 months, stroke participants had higher CCI scores than controls and were more likely to have a history of T2DM, hypertension, smoking and atrial fibrillation (Table 1). At 3 years, stroke participants had more atrial fibrillation than controls.

Cognitive impairment at 3 months was associated with greater age (71.8±8.9 years CI vs 64.6±12.4 CN), higher CCI (4 CI vs 3 CN) and hypertension (80% CI vs 46% CN; Table 2).

### Cognitive

Control participants had significantly greater years of education and higher NART-FSIQ scores compared to stroke participants (Table 1). No associations were observed between years of education and NART-FSIQ and cognitive impairment in stroke participants (Table 2).

### Imaging and stroke characteristics

Stroke participants had smaller HV than controls at 3 months (3862±490mm^3^ vs 4032±343 mm^3^, Table 1) and higher WMH volumes than controls at both time-points (e.g., 3-month median stroke=866 mm^3^, controls=684 mm^3^, p=0.0001; Table 1; Supplemental Tables). Cognitively impaired stroke participants had smaller TBV (CN=1119±108cm^3^, CI=1026±110cm^3^) and HV (CN=4047±394 mm^3^, CI=3589±360mm^3^) and larger stroke lesion volumes (CN=1181mm^3^, CI=5899mm^3^) but not WMH volume (median CN=1572 mm^3^, CI=2069 mm^3^). Cognitive impairment was not associated with admission NIHSS, prior stroke, or side of stroke (Table 2). We observed more large-artery embolic and less lacunar strokes in the CI group.

### Primary outcome: change in TBV

Figure 1 and Table 3 demonstrate that both groups lost TBV between 3-months and 3-years, but TBV loss in stroke participants (n=93, 26 women, 66.7±12 years) was significantly greater than controls (n=39, 15 women, 68.7± 6.7 years): stroke mean 20.31cm^3^±SD14.8cm^3^; controls mean 14.22 cm^3^±SD13.21cm^3^; adjusted mean difference 7.88 95% CI [2.84, 12.91] p-value 0.002. This was confirmed on sensitivity analysis (n=122 stroke, n=40 controls, p-value for group-by-time interaction 0.002).

**Figure 1.**
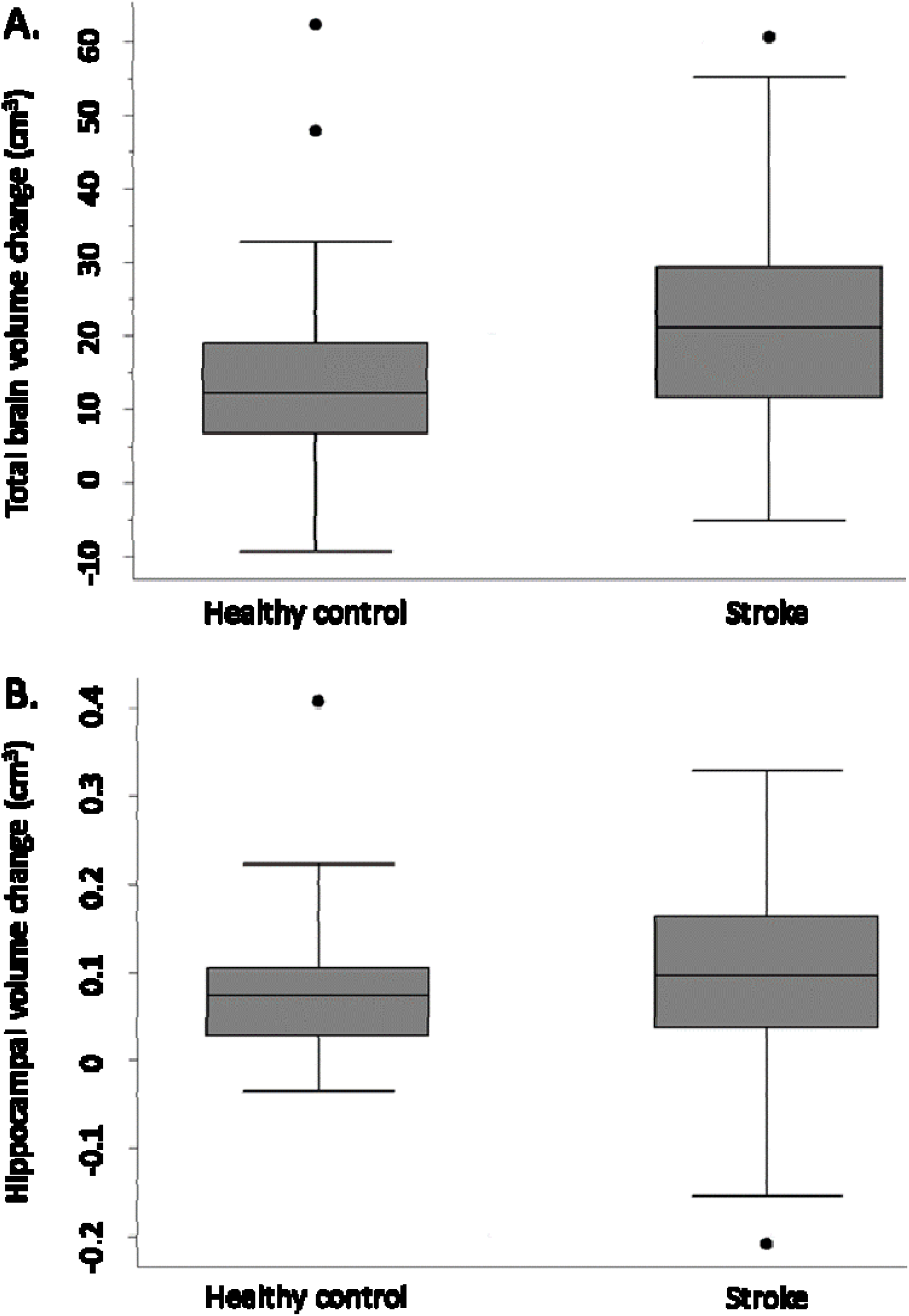
Total brain and hippocampal volume change stroke vs controls. Total brain volume (Panel A) and hippocampal volume (Panel B) change between 3-months and 3-years in stroke patients and healthy controls. Y-axis=change in brain volume (cm^3^). Positive values represent decline in brain volumes: i.e., greater brain volume loss. Midline=median brain volume change. X-axis: n=93 stroke, n=39 healthy controls.

**Table 3.** Primary and secondary hypotheses results: linear regression analyses. A: Primary outcome difference between stroke and control TBV change (TBVΔ); B: Secondary outcome 2: Difference between stroke and control HV change (HVΔ); C: Difference between TBVΔ in CN vs CI stroke participants; D: Difference between HVΔ in CN vs CI stroke participants. All volume values mean(SD) in cm^3^

| **Brain volume comparison between stroke and control participants** |  |  |  |  |
| --- | --- | --- | --- | --- |
| **Outcome** | **Stroke** | **Control** | **Adjusted mean difference [95%CI]** | **p-value** |
| **A. TBVΔ** | 20.31(14.84) | 14.22(13.21) | 7.88<br>[2.84,12.91] | 0.002 |
| **B. HVΔ** | 0.09(0.09) | 0.08(0.08) | 0.02<br>[-0.02,0.05] | 0.32 |
| **Comparison between CN and CI stroke participants** |  |  |  |  |
|  | **CN** | **CI** | **Adjusted mean difference [95%CI]** | **p-value** |
| **C. TBVΔ** | 19.63(13.84) | 30.67(14.18) | 10.42<br>[3.04,17.80] | 0.006 |
| **D: HVΔ** | 0.09(0.10) | 0.11(0.09) | 0.01<br>[-0.04,0.06] | 0.61 |

### Secondary outcome 1

Figure 2 shows that TBV decline was greater in the CI group (mean=30.67±SD14.18 cm^3^) relative to the CN group (mean=19.63 cm^3^±13.84 cm^3^); adjusted mean difference 10.42; 95% CI [3.04, 17.80], p-value=0.006; Figure 2), confirmed on sensitivity analysis (CN n=80, CI n=41, p-value for group-by-time interaction 0.001).

**Figure 2.**
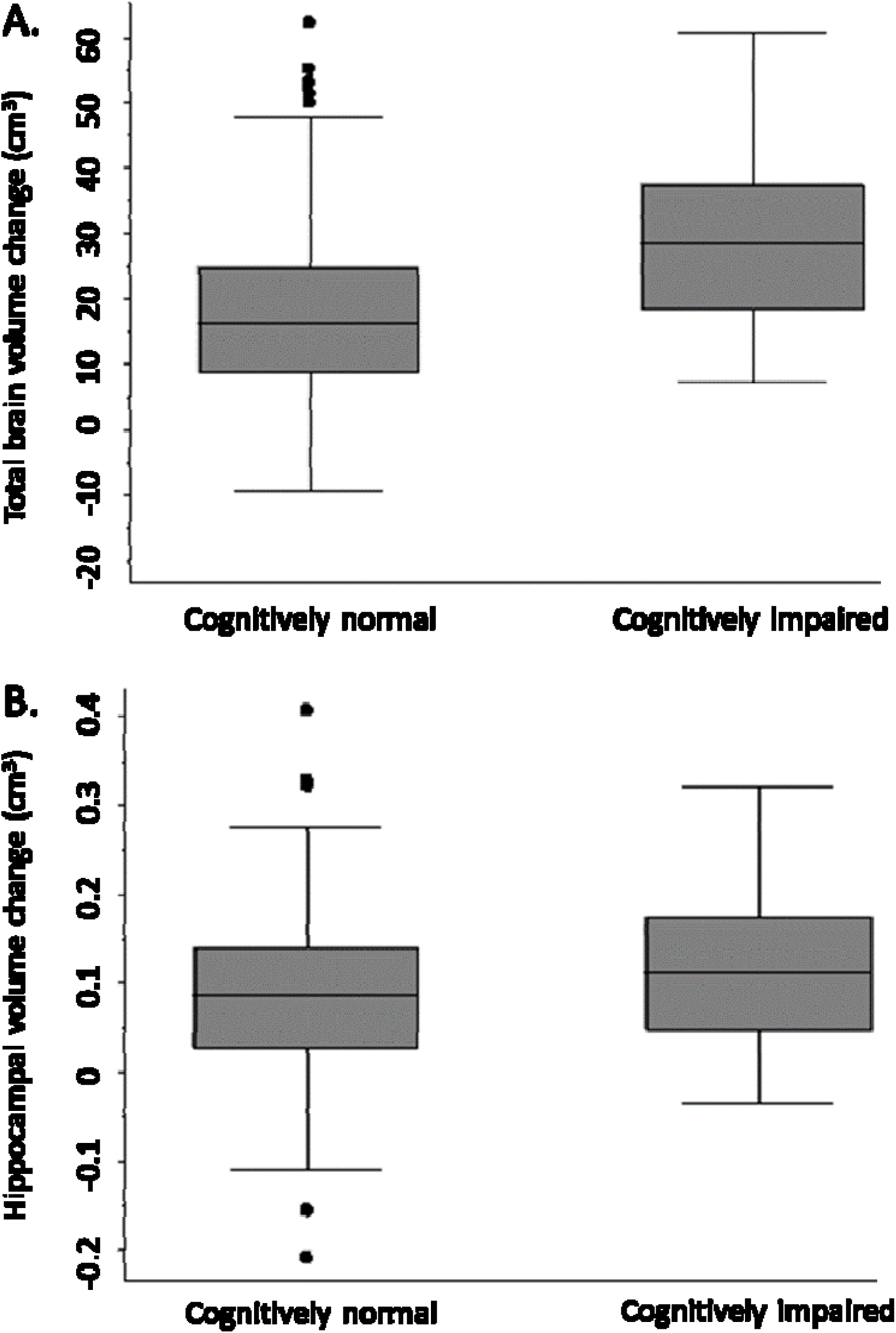
Total brain and hippocampal volume change cognitively normal vs impaired. Total brain volume (Panel A) and hippocampal volume (Panel B) change between 3-months and 3-years post-stroke in CN and CI stroke groups determined at 3-months. Y-axis=change in brain volume(cm^3^). Positive values represent decline in brain volume, negative values represent an increase in brain volume. Midline=median brain volume change.

### Secondary outcomes 2 and 3

HV change was comparable in both stroke and control participants (stroke n=93, controls n=39, complete case analysis adjusted mean difference 0.02, 95% CI [-0.02, 0.05], p=0.32; sensitivity analysis: n=40 controls, n=122 strokes, p-value for group-by-time interaction 0.26). No significant difference in HV change was observed between CN and CI stroke participants (complete case analysis adjusted mean difference 0.01 [-0.04, 0.06], p-value =0.61; sensitivity analysis: CN n=80 CI, n=41, p-value for group-by-time interaction 0.41).

## DISCUSSION

We report the primary and secondary outcomes from CANVAS, a prospective cohort study of ischaemic stroke patients compared to age- and sex-matched controls where all participants were cognitively normal at recruitment and all ischaemic stroke subtypes were included. We found that TBV loss in stroke participants over 3 years was significantly greater than controls. Further, TBV loss over 3 years was greater in stroke patients who were cognitively impaired at 3 months compared to those without cognitive impairment. We did not find these associations for hippocampal volume loss, although cognitively impaired participants had smaller hippocampi than those who were cognitively normal.

No participants had a history of cognitive decline prior to study entry. One-third of our stroke participants were cognitively impaired 3 months after stroke, and 26% were impaired at 3 years, but more participants who were cognitively impaired at 3 months died or did not attend the 3-year visit. In addition, we found an association with higher BMI and incomplete data sets. Cognitively impaired stroke participants at 3 months had smaller brain volumes, larger stroke lesion volumes, more vascular risk factors, and higher CCI scores, but not WMH volume, than those who were cognitively normal.

Hippocampal atrophy is strongly associated with vascular risk factors. We have previously reported smaller hippocampi in stroke than control participants at baseline^8^, and shown that atrophy rates are greater in 3 months after stroke^10^ than the ensuing 9 months. However, we did not find that stroke hippocampal atrophy rates were greater over 3 years than controls, nor did we find an association with cognitive impairment. This could be because most of the hippocampal atrophy had already occurred – a potential floor effect. Many authors have proposed that incipient Alzheimer’s pathology is contributing to the hippocampal atrophy, and that pre-existing protein deposition leads to the observed atrophy. However, this has not been borne out on PET amyloid imaging studies of stroke^30, 31^, nor did we find an association between hippocampal volume and amyloid status in a small PET amyloid sub-study at 3 years^32^. Vascular risk factors are increasingly considered as primary drivers of hippocampal degeneration ^9, 10^.

Also, the lack of association between hippocampal atrophy and post-stroke cognitive impairment may not be surprising when viewed in the clinical context of vascular cognitive deficits. We did not use paired associate measures as our verbal memory test, which are more sensitive to hippocampal dysfunction. Vascular cognitive impairment is not characterised by the primary amnestic deficits seen in clinical Alzheimer’s dementia. Rather, it is characterised by slowed speed of processing, memory retrieval problems, behavioral, attention and executive dysfunction, which are dependent on normal thalamic and frontal white matter tract and cortical function. Thalamic dysfunction has been implicated in the clinical impairments seen after stroke, with ipsi- and contralesional thalamic atrophy over the first year reported^10^. In addition, we have previously reported that hippocampal subfield atrophy correlated better with verbal memory impairment in this cohort, suggesting that whole hippocampal atrophy may not be the metric of choice in post-stroke cognitive impairment^33^.

The observed reduction in TBV in stroke survivors and its strong association with cognitive impairment at 3 months are novel. Imaging measures of global brain volume necessarily capture both grey and white matter atrophy. The latter is often underestimated in histopathological studies, as white matter atrophy is hard to estimate histologically. Advances in neuroimaging have allowed us to chart white matter loss after stroke^9^. The frontal lobes are especially vulnerable to vascular brain burden^34^, and attention and executive function are dependent on the integrity of these frontal networks. We have demonstrated degeneration of the frontal distributed brain networks subserving attention and executive function one year after stroke ^34, 35^. The deficits associated with vascular cognitive impairments could be posited as disconnection syndromes, arising both from the stroke lesions and from concomitant white matter disease.

It is likely that there are multiple mechanisms underlying this white matter degeneration in stroke populations, but we know that brain infarction triggers extensive, pervasive neuroinflammation^36^. Microglial activation occurs within brain networks involving the infarct, including the thalami bilaterally^36^. These sustained inflammatory responses may contribute to the Wallerian degeneration in tracts connected to the stroke. This ongoing grey and white matter degeneration^9, 10^, captured in our study by the increased TBV loss seen in the stroke group, may manifest clinically as cognitive decline.

Study strengths include the strongly positive primary hypothesis, and our prospective, observational control-cohort design with identical study visits, imaging protocols and cognitive tasks for all participants. Our high-quality imaging, multi-domain approach to cognition, expected attrition rate and documented missing data are also strengths. Study weaknesses include the small numbers compared to large community-based studies, incomplete data sets associated with older age, relatively mild, mainly male, stroke participants, and our healthier, more educated control participants – a common problem with volunteer bias, especially imaging studies^37^. It can be very difficult to match vascular risk factors in stroke-free controls, as these same risk factors obviously increase their chances of incidental or concomitant stroke. Unlike some other, larger studies, our primary objective was not to examine determinants of post-stroke dementia, and therefore we were not powered to look at predictors of cognitive decline and dementia at 3 years, but plan to pool these data with larger prospective studies including stroke participants alone.

## CONCLUSIONS

Ischaemic stroke survivors exhibit greater neurodegeneration compared to stroke-free controls. Atrophy is greater in those who are cognitively impaired early after stroke. The coupling of early cognitive impairment and greater subsequent atrophy likely reflects the combined impacts of stroke and vascular brain burden. Our results help to disentangle the complex interactions between incipient cognitive decline, the cumulative impact of vascular risk factors, and the effect of the super-added stroke lesion. These brain atrophy rates can be used for future intervention trials to reduce post-stroke neurodegeneration.

## Supporting information

STROBE_checklist

## Data Availability

The data collected for this study is available on request to the authors. The study protocol was published in 2014 and is freely available. Data requests should be sent to the corresponding author and will be evaluated and approved after consultation with CANVAS chief investigators. One-year data (not part of this study) have already been shared with the ENIGMA Stroke Recovery consortium http://enigma.ini.usc.edu/ongoing/enigma-stroke-recovery/. We have committed to sharing CANVAS data with the STROKOG consortium. CANVAS also has a 5-year follow-up timepoint (not presented here). It is a requirement of the funding for this timepoint that it will be made publicly available once complete (expected completion and compilation late 2021 due to pandemic delays). This will include de-identified participant data including MRI data. Data will be shared at reasonable request following review by the CANVAS data steering committee.

## Registration

**http://www.clinicaltrials.gov NCT02205424**

## Acknowledgements

The authors thank the University of Melbourne Victorian Life Sciences Computation Initiative, Florey Node National Imaging Facility, Melbourne Brain Centre radiographers, and all our participants, who so generously contributed their time to the study.

## Supplemental Material

- Expanded Materials & Methods
- Tables I-III
- Figures I – II

## Non-standard Abbreviations and Acronyms

CANVAS: Cognition And Neocortical Volume After Stroke
CCI: Charlson Comorbidity Index
CDR: Clinical Dementia Rating
CI: cognitively impaired
CN: cognitively normal
HV: hippocampal volume
IQCODE: Informant Questionnaire on Cognitive Decline in the Elderly Short Form
NART: National Adult Reading Test
TBV: total brain volume

