## Supplemental_Materials for "Increased brain atrophy over 3 years in stroke survivors is linked to early post-stroke cognitive impairment"

**SUPPLEMENTAL MATERIAL**

- **Expanded Materials & Methods**
- **Online Tables I-III**
- **Online Figures I – II**
- **References**

**Methods**

**Details of imaging sequences and analytic approach**

The MR images for the presented analyses were acquired using a T1-weighted 3D magnetisation-prepared rapid gradient (MPRAGE) sequence with the following parameters: 160 sagittal slices, repetition time (RT)=1900 ms, echo time (TE)=2.6 ms, inversion time (TI)=900 ms, flip angle=9°, matrix size=256×256, slice thickness=1 mm, voxel size=1×1×1 mm^3^ isotropic. High-resolution, 3D sampling with application-optimised contrasts using different flip angle evolution fluid-attenuated inversion recovery (FLAIR) images were also acquired for white matter hyperintensity estimation and stroke lesion tracing: 160 sagittal slices, 1 mm thick, RT=6000 ms, TE= 380 ms, 120° flip angle, and 256x256 acquisition matrix. Diffusion weighted imaging was also performed, with a shorter, “clinical” sequence to examine for the presence of recent (last 10-14 days) interval stroke.

Stroke lesions were traced by our imaging analyst (MSK) and cross-checked by a stroke neurologist (AB). Stroke lesion sites were also cross-checked with acute inpatient imaging, particularly their DWI lesion site on acute MRI where available, as white matter infarcts were sometimes difficult to delineate on the 3-month scan in patients with severe white matter disease.

The FSL software package was used to generate binary lesion masks and compute lesion volumes (<https://protect-au.mimecast.com/s/aPrcCL7rxDsRXnPPPHBdMbu?domain=fsl.fmrib.ox.ac.uk/>).

The probability maps of white matter hyperintensities (WMH) were obtained from FLAIR images using the lesion prediction algorithm (LPA)^1^ from the lesion segmentation toolbox (LST) (https://applied-statistics.de/lst.html) included in SPM12. The WMH maps were first eroded by masks of stroke lesions before WMH volumes were computed using ‘fslstats’ and a threshold of 0.5.

**Cognitive Outcome Evaluation Committee**

Outcome evaluation committee meetings were convened for allocation of cognitive status. The committee included cognitive and stroke neurologists (AB, DD), clinical (JB) and research neuropsychologists (EW, TC), and research psychologists (LB). The committee met in person at pre-agreed times on 4 occasions after the 3-year assessment was complete. Judgements regarding impact on functional independence were specifically directed to the influence of their cognitive impairment not their stroke-related disability if residual.

De-identified information from their structured clinical interview (e.g., information regarding relevant recent psychosocial stressors, functional status), standardised cognitive z-scores, mood scores, and the Global CDR Score were presented to our Outcome Evaluation Committee. Participants were assigned their status of *normal cognition (CN), cognitively impaired (CI),* or *dementia,* in these panel meetings. We also allowed the assignment of *unclassifiable* if the participant was unable to complete sufficient cognitive testing and/or no history was obtained from an informant.

A participant was assigned *cognitively normal* (CN) if z-scores in all cognitive domains were within accepted age- and years of education-adjusted norms and there was no evidence of functional decline due to cognitive impairment (i.e., activities of daily living (ADLs) were unaffected).

A participant was judged to be *CI* if (1) the z-score for at least one cognitive domain was lower than -1.5, and (2) there was no evidence of functional decline due to cognitive impairment (ADLs were unaffected).

A participant was judged to have *dementia* if (1) z-scores for two or more cognitive domains were lower than -1.5 and (2) there was evidence of functional decline (i.e., significant impact on activities of daily living between time-points due to cognitive impairment, as demonstrated by (a) discussions with next-of-kin and the Short-Form IQCODE, and/or (b) CDR score of 1 or greater.

**Sample size calculation**

We calculated our sample size to include sufficient participants for both the primary outcome and secondary outcome 1. The primary outcome required a direct comparison of two groups, stroke versus control participants. We estimated that stroke patients would lose brain volume at around twice the rate of controls based on pilot^2^ and published data^3, 4^: 1% TBV reduction each year over 3 years. Healthy elderly controls lose brain volume around 0.5%/year^4^, but rates are much higher in brain disease (stroke 2%/year^3^, dementia s 2-5%/year^4^). We estimated 35 participants per group yielding power of 0.8 assuming two-sided alpha of 0.05.

For the secondary outcome 1, we designed the sample of stroke participants to include an expected prevalence of cognitive impairment of 30% at 3 months^5^. We used an ANCOVA method to estimate sample size for two samples with repeated measures (CN versus CI stroke in a ratio of 2:1), including a correlation score between baseline and follow-up. Using an alpha level of 0.05 (two-sided), power=0.8 and a conservative correlation of 0.1, we estimated that 108 participants would be required (36 CI and 72 CN). We predicted 20% attrition due to death or non-participation, meaning total 135 stroke and 40 control participants.

**Data availability statement**

The data collected for this study is available on request to the authors. The study protocol was published in 2014 and is freely available^6^. Data requests should be sent to the corresponding author and will be evaluated and approved after consultation with CANVAS chief investigators.

**Supplemental Tables**

**Supplemental Table I**

**Characteristics of participants with complete data included in the 3-year analyses.**

Demographic, clinical, imaging, cognitive and stroke characteristics comparing characteristics of stroke patients and healthy controls at 3 years. Original stroke characteristics included for all participants included at 3 years. *APOE*=apolipoprotein E; WMH=white matter hyperintensity; NART-FSIQ=National Adult Reading Test estimated Full-Scale IQ; aNIHSS=admission National Institute of Health Stroke Scale.

|  | **Stroke** | | **Control** | |  |
| --- | --- | --- | --- | --- | --- |
|  | N |  | N |  | *p* |
| **Sociodemographic** | | | | | |
| Age, years,  M ± SD | 93 | 66.72 ± 11.97 | 39 | 68.72 ± 6.68 | 0.33 |
| Men, no. (%) | 93 | 67 (72%) | 39 | 24 (61.5%) | 0.30 |
| **Clinical** | | | | | |
| *APOE ε4*, no. (%) | 90 | 17 (18.9%) | 38 | 4 (10.5%) | 0.30 |
| Depression, no. (%) | 93 | 9 (9.7%) | 39 | 4 (10.3%) | >0.99 |
| Charlson Comorbidity Scores, Md (Q1, Q3) | 93 | 3 (2, 4) | 39 | 3 (2, 3) | 0.26 |
| Hypertension, no. (%) | 93 | 50 (53.8%) | 39 | 17 (43.6%) | 0.34 |
| Hypercholesterolaemia, no (%) | 93 | 38 (40.9%) | 39 | 14 (35.9%) | 0.70 |
| Type 2 diabetes mellitus, no. (%) | 93 | 20 (21.5%) | 39 | 4 (10.3%) | 0.15 |
| Atrial fibrillation, no (%) | 93 | 18 (19.4%) | 39 | 1 (2.6%) | 0.013 |
| Smoking, pack years,  Md (Q1, Q3) | 90 | 1 (0, 13) | 39 | 0 (0, 3) | 0.92 |
| Obesity, no. (%) | 93 | 30.1% | 39 | 7 (18%) | 0.20 |
| **Imaging** | | | | | |
| Days to scan (Md, Q1, Q3) | 93 | 1106 (1096, 1128) | 39 | 1111 (1096, 1121) | 0.70 |
| Total brain volume (cm^3^, M±SD) | 93 | 1070 (116) | 39 | 1096 (80) | 0.15 |
| Hippocampal volume (mm^3^, M±SD) | 93 | 3817 (457) | 39 | 3947 (357) | 0.084 |
| Stroke lesion volume  (mm^3^, Md, Q1, Q3) | 93 | 2014 (341, 11375) |  | - |  |
| WMH Volume (mm^3^, Md, Q1, Q3) | 93 | 3040 (970, 7284) | 39 | 1189 (523, 2936) | 0.0027 |
| **Cognitive** | | | | | |
| Education, years,  Md (Q1, Q3) | 93 | 12 (10, 15) | 39 | 17 (11, 18) | 0.001 |
| NART-FSIQ, Md (Q1, Q3) | 90 | 112 (103.1, 120.9) | 39 | 120.9 (111.1, 124.9) | <0.001 |
| **Stroke Characteristics** | | | | | |
| Previous stroke, no. (%) | 102 | 15 (15) |  | - | - |
| Thrombolysis, no. (%) | 102 | 14 (14) |  | - | - |
| aNIHSS score, Md (Q1, Q3) | 102 | 0 (0, 1) |  | - | - |
| aNIHSS severity, mild (0-7), no. (%) | 102 | 102 (100) |  | - | - |
| aNIHSS score, Md (Q1, Q3) | 102 | 2 (1, 4) |  | - | - |
| aNIHSS severity, mild (0-7), no. (%) | 102 | 95 (93.1) |  | - | - |
| **Stroke side, no. (%)** | 102 |  |  |  |  |
| Right |  | 59 (58) |  | - | - |
| Left |  | 40 (39) |  | - | - |
| Bilateral |  | 3 (3) |  | - | - |
| **Oxfordshire, no. (%)** | 102 |  |  |  |  |
| Lacunar |  | 14 (14) |  | - | - |
| Posterior |  | 33 (32) |  | - | - |
| Partial anterior |  | 54 (53) |  | - | - |
| Total anterior |  | 1 (1) |  | - | - |
| **TOAST, no. (%)** | 102 |  |  |  |  |
| Large artery |  | 17 (17) |  | - | - |
| Cardioembolic |  | 27 (26) |  | - | - |
| Small-vessel occlusion |  | 16 (16) |  | - | - |
| Other determined |  | 6 (6) |  | - | - |
| Undetermined |  | 36 (35) |  | - | - |

**Supplemental Table II**

**Comparison of characteristics of stroke participants with complete and incomplete study data at 3 months**.

We have compared sociodemographic, clinical, and cognitive data for all those participants with complete data sets at 3 months and those who were excluded due to non-attendance, incomplete assessments, or non-evaluable scans. P-values are included for interest however given low numbers may not be valid. Imaging details are not included as these data were not available. *APOE*=apolipoprotein E; WMH=white matter hyperintensity; NART-FSIQ=National Adult Reading Test estimated Full-Scale IQ; aNIHSS=admission National Institute of Health Stroke Scale.

|  | **Participants with 3- month measure** | | **Participants without 3-month measures** | |  |
| --- | --- | --- | --- | --- | --- |
|  | N |  | N |  | *p* |
| **Sociodemographic** | | | | | |
| Age, years,  M ± SD | 122 | 67.8 (11.8) | 13 | 71.2 (20.8) | 0.57 |
| Men, no. (%) | 122 | 84 (68.9%) | 13 | 10 (76.9%) | 0.75 |
| **Clinical** | | | | | |
| *APOE ε4*, no. (%) | 104 | 20 (19.2%) | 9 | 1 (11.1%) | >0.99 |
| Depression, no. (%) | 122 | 11 (9.0%) | 7 | 2 (28.6%) | 0.15 |
| Charlson Comorbidity Scores, Md (Q1, Q3) | 122 | 3 (2, 5) | 4 | 4 (3.5, 5.5) | 0.23 |
| Hypertension, no. (%) | 122 | 78 (63.9%) | 13 | 7 (53.9%) | 0.55 |
| Hypercholesterolaemia, no (%) | 122 | 53 (43.4%) | 13 | 9 (69.2%) | 0.087 |
| Type 2 diabetes mellitus, no. (%) | 122 | 31 (25.4%) | 13 | 3 (23.1%) | >0.99 |
| Atrial fibrillation, no (%) | 122 | 27 (22.1%) | 13 | 6 (46.2%) | 0.084 |
| Smoking, pack years,  Md (Q1, Q3) | 120 | 1 (0, 17) | 11 | 0 (0, 27.8) | 0.99 |
| Obesity, no. (%) | 122 | 34 (27.9%) | 13 | 11 (84.6%) | <0.001 |
| **Cognitive** | | | | | |
| Education, years,  Md (Q1, Q3) | 122 | 12 (10, 15) | 13 | 11 (9.8, 12.3) | 0.21 |
| NART-FSIQ, Md (Q1, Q3) | 112 | 111 (103, 121) | 5 | 105 (103, 109) | 0.29 |
| **Stroke Characteristics** | | | | | |
| Previous stroke, no. (%) | 122 | 17 (13.9%) | 13 | 4 (30.8%) | 0.12 |
| Thrombolysis, no. (%) | 122 | 15 (12.3%) | 13 | 2 (15.4%) | 0.67 |
| aNIHSS score, Md (Q1, Q3) | 122 | 2 (1, 4) | 13 | 2 (1.75, 3.25) | 0.86 |
| aNIHSS severity, mild (0-7), no. (%) | 122 | 114 (93.4%) | 13 | 13 (100%) | >0.99 |
| mRS score at assessment, Md (Q1, Q3) | 120 | 1 (1, 2) | 4 | 2 (1, 2) | 0.37 |
| mRS severity, mild (0-1), no. (%) | 120 | 84 (70.0%) | 4 | 1 (25.0%) | 0.092 |

**Supplemental Table III**

**Comparison of characteristics of stroke participants with complete and incomplete study data at 3 years.**

We have compared sociodemographic, clinical, and cognitive data for all those participants with complete data sets at 3 years and those who were excluded due to non-attendance, incomplete assessments, or non-evaluable scans.

|  | **Participants with 3- year measures** | | **Participants without 3-year measures** | |  |
| --- | --- | --- | --- | --- | --- |
|  | N |  | N |  | *p* |
| **Sociodemographic** | | | | | |
| Age, years,  M ± SD | 93 | 69.5 (12.0) | 29 | 75.7 (10.4) | 0.01 |
| Men, no. (%) | 93 | 67 (72.0%) | 29 | 17 (58.6%) | 0.18 |
| **Clinical** | | | | | |
| APOE ε4, no. (%) | 86 | 17 (19.8%) | 18 | 3 (16.7%) | >0.99 |
| Depression, no. (%) | 93 | 9 (9.7%) | 29 | 2 (6.9%) | >0.99 |
| Charlson Comorbidity Scores (CCI), Md (Q1, Q3) | 93 | 3 (2, 5) | 4 | 4.5 (3.5, 5.5) | 0.19 |
| Hypertension, no. (%) | 93 | 52 (55.9%) | 29 | 26 (89.7%) | <0.001 |
| Hypercholesterolaemia, no (%) | 93 | 38 (40.9%) | 29 | 15 (51.7%) | 0.39 |
| Type 2 diabetes mellitus, no. (%) | 93 | 20 (21.5%) | 29 | 11 (37.9%) | 0.090 |
| Atrial fibrillation, no (%) | 93 | 17 (18.3%) | 29 | 10 (34.5%) | 0.077 |
| Smoking, pack years,  Md (Q1, Q3) | 91 | 1 (0, 14.5) | 29 | 3 (0, 24.8) | 0.45 |
| Obesity, no. (%) | 93 | 27 (29.0%) | 29 | 26 (89.7%) | <0.001 |
| **Cognitive** | | | | | |
| Education, years,  Md (Q1, Q3) | 93 | 12 (10, 15.3) | 29 | 12 (10, 15) | 0.41 |
| NART-FSIQ, Md (Q1, Q3) | 90 | 112 (103, 121 | 22 | 110 (103, 118) | 0.80 |
| **Stroke Characteristics** | | | | | |
| Previous stroke, no. (%) | 93 | 10 (10.8%) | 29 | 7 (24.1%) | 0.12 |
| Thrombolysis, no. (%) | 93 | 13 (14.0%) | 29 | 2 (6.9%) | 0.52 |
| NIHSS score, Md (Q1, Q3) | 93 | 0 (0, 1) | 4 | 0 (0, 1) | 0.98 |
| NIHSS severity, mild (0-7), no. (%) | 93 | 93 (100%) | 4 | 4 (100%) | >0.99 |
| mRS score at assessment, Md (Q1, Q3) | 93 | 1 (0, 1) | 4 | 1 (0.5, 1.5) | 0.73 |
| mRS severity, mild (0-1), no. (%) | 93 | 74 (79.6%) | 4 | 3 (75.0%) | 1.0 |

**Supplemental Figures**

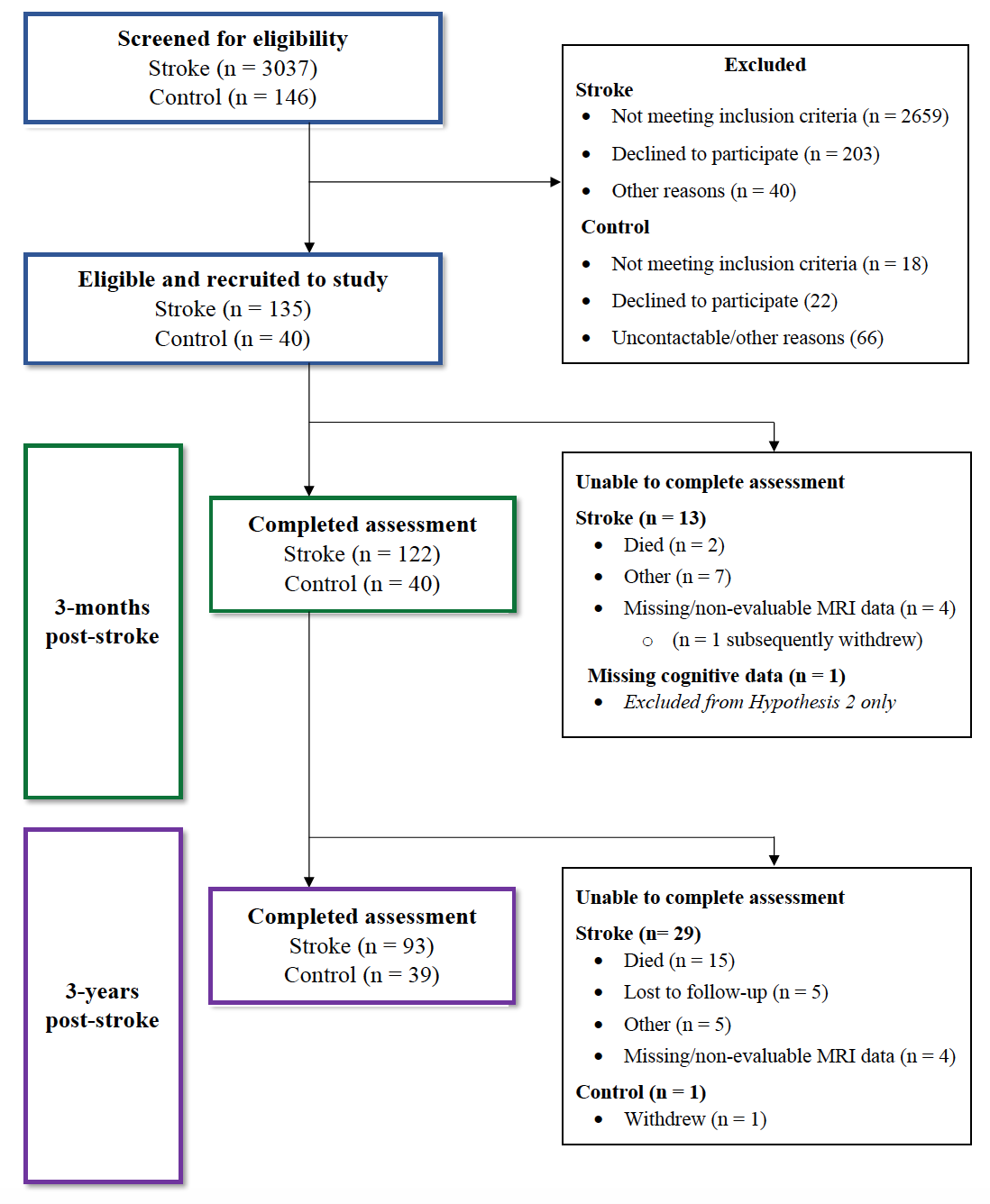

**Supplemental Figure 1**

**CONSORT-style flow-chart of screening, recruitment, and retention over the study.**

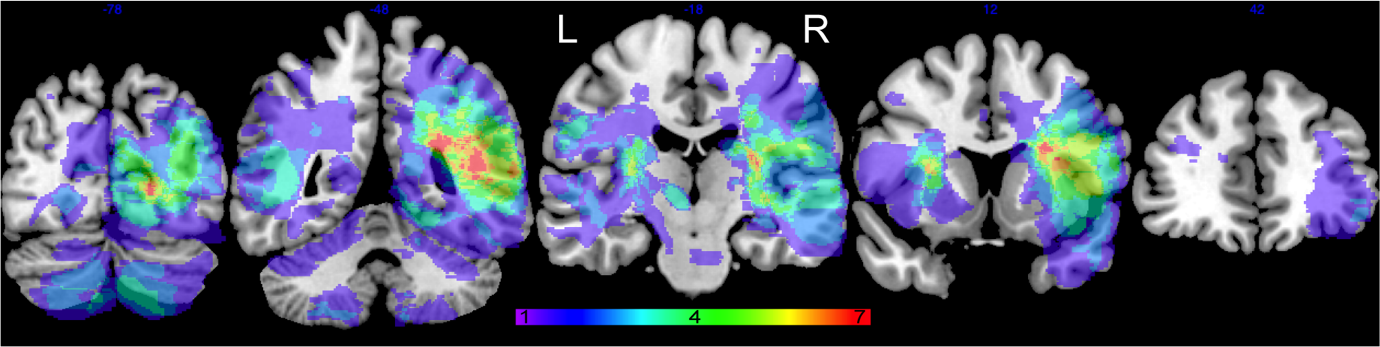

**Supplemental Figure 2**

**Stroke lesion map.** Lesions traced on 3-month MRI data for the 93 participants with 3-­year data. The map shows an overlay of lesion sites: warmer colors indicate greater overlap (e.g., from 1=purple to 7=red) of lesions. Lesion maps were prepared using MRIcron software.
